# COVID-19 mRNA-vaccination and all-cause mortality in the adult population in Norway during 2021-2023: a population-based cohort study

**DOI:** 10.1101/2024.12.15.24319058

**Authors:** Jesper Dahl, German Tapia, Håkon Bøås, Inger Johanne Landsjøåsen Bakken, Hanne Løvdal Gulseth

## Abstract

**Introduction:** Most countries worldwide have experienced excess mortality that coincides temporally with the COVID-19 mass vaccination campaigns. This has led to speculation on the potential long-term effects of the vaccines on mortality risk.

**Methods:** The study was designed as a retrospective cohort study, and included all individuals aged ≥18 years living in Norway during January 1, 2021, through December 31, 2023. Individuals were categorized as either unvaccinated (received no doses), partially vaccinated (received one or two doses) or fully vaccinated (received three or more doses) from the date of vaccination and onwards. Age-stratified Poisson models were used to estimate incidence rate ratios of death (all causes) between vaccination groups, adjusting for sex, calendar time, county of residence and risk group status (nursing home resident or preexisting condition with increased risk of severe COVID-19).

**Results:** The study included 4 645 910 individuals (49.8% women) with 132 963 deaths during follow-up. There was a higher proportion of individuals that were part of a risk group among fully vaccinated individuals compared to unvaccinated individuals in all age groups, and a lower unadjusted rate of death: 51.5 vs 73.6 per 100 000 person years (py) among individuals aged 18-44 years, 295.1 vs 405.3 per 100 000 py among 45-64 years, and 3620.2 vs 4783.8 per 100 000 py among 65 years or older. The adjusted IRR of death for the same age groups were 0.42 (95% CI 0.38-0.47), 0.39 (95% CI 0.37-0.41) and 0.42 (95% CI 0.41-0.43), respectively. The differences in rate of death between vaccination groups were larger among men and peaked during 2022.

**Conclusion:** Vaccinated individuals had a lower rate of all-cause death during 2021-2023 in Norway.

**Key messages:** In Norway, as in many Western countries, there has been an excess mortality during 2021-2023, surpassing the numerical count of COVID-19-associated deaths. The excess mortality coincides temporally with the COVID-19 mass vaccination campaigns, but longitudinal data regarding mortality post-vaccination remain sparse. Using real-time national health registry data, we estimate the risk of all-cause mortality by vaccination status in the total adult population in Norway during 2021-2023 and demonstrate a lower rate of all-cause death among vaccinated individuals.

## Introduction

Most countries worldwide have experienced increased all-cause mortality during the covid-19 pandemic. This excess mortality has generally been attributed to surges in covid-19 cases(1), but there are still lingering questions as to the indirect effects of the pandemic, such as disruption of health services, on the observed mortality rates(2). Due to the proximity of the widespread implementation of covid-19 vaccination programmes to the observed excess mortality, there has also been speculation as to whether vaccinations can have contributed to the increased mortality risk(3). This has generally been refuted by studies on vaccination effect, which have shown reduced mortality rates among vaccinated individuals (4), and studies presenting no increased mortality in vaccinated individuals(5–9). We have earlier investigated short-term vaccination in the elderly(10). However, follow-up time in these studies is generally limited to <1 year, which has left room for speculations of a potential long-term effect of vaccination on mortality risk. To address these concerns, we aimed to present all-cause mortality rates by vaccination status throughout all periods with observed excess mortality in Norway during the covid-19 pandemic, while accounting for existing medical conditions with increased risk of severe COVID-19.

## Methods

### Study design

The study was designed as a retrospective cohort study and included all individuals with a national identity number (all permanent residents) aged 18 year or older living in Norway during January 1, 2021, through December 31, 2023. Date of birth, immigration, emigration and death was retrieved from the National Population Register. Each individual could enter follow-up at any point of the study period upon attaining 18 years of age and/or immigrating, and was censored at emigration, death or end of follow-up (December 31, 2023).

### Data Source

The Emergency preparedness register for COVID-19 (Beredt C19) was established according to the Health Preparedness Act §2-4 to assess risk and implement measures during the COVID-19 pandemic(11). Beredt C19 receives registry data from mandatory Norwegian health registries and other relevant data sources at varying time intervals, commonly on a weekly basis.

For the current study, we used data originating from the Norwegian Immunisation Registry (SYSVAK), the National Population Register, the Norwegian Patient Registry (NPR) and the Norwegian Registry for Primary Health Care (NRPC), as described in detail below.

All data linkage and analyses were conducted within Beredt C19. Individual-level registry information was linked using the unique personal identification number (pseudonymized) given to every resident at birth or immigration.

### Exposure - Vaccination

All covid-19 vaccinations in Norway are mandatorily reported to SYSVAK. Using the type of vaccine and date of vaccination we categorized all person time into “received no doses” (from here on referred to as “unvaccinated”), “received one or two doses of covid-19 vaccination” (from here on referred to as “partially vaccinated”) or “received 3 or more doses of covid-19 vaccination” (from here on referred to as “fully vaccinated”). Each individual was counted as vaccinated from the date of vaccination and onwards, and only changed groups upon receiving a new relevant dose. Only doses with mRNA vaccines were included in the current study: BNT03/Comirnaty, CBA01/Omicron BA.1, CBA45/Comirnaty Omicron BA.4-5, CBB15/Comirnaty Omicron XBB.1.5, MOD03/ Spikevax, SBA01/ Spikevax Omicron BA.1, SBA45/Spikevax Omicron BA.4-5(12). These vaccine types accounted for 98.7% of all covid-19 vaccine doses registered in SYSVAK up until December 31, 2023. If an individual received a dose with another type of covid-19 vaccination, they were censored from the analysis on the day prior to this vaccination.

### Outcome – All-cause death

Death from any cause was considered the main outcome in each analysis. Date of death was retrieved from the National Population Register.

### Covariates

Each analysis was stratified by age group (18-44 years, 45-64 years and 65 years or older) and included the following covariates: Sex, county of residence, calendar time (quarter and year) and medical risk group at baseline. Each analysis is also presented sex-stratified, with sex being removed as a covariate from the models in question.

Vaccine recommendations in Norway during the study period were in large part based on age, and the given age groups were chosen to reflect these differing recommendations.

Information on sex and county of residence was retrieved from the National Population Register and included as categorical variables.

Information on age was also retrieved from the National Population Register and was handled as a time-varying categorical variable (“18-44 years”, “45-64 years” and “65 years or older”), meaning that each individual could change age-groups during follow-up.

Calendar time (quarter and year) was included as a time-varying covariate. Q1 was defined as January 1 through March 31, Q2 as April 1 through June 30, Q3 as July 1 through September 30, and Q4 as October 1 through December 31.

Medical risk group was included as a dichotomous covariate (yes/no). It was defined using the NPR and the NRPHC(13). Any individual on short- or long-term nursing home stay or with a preexisting, predefined risk condition at start of follow-up was considered as being in the risk group. The definition of risk conditions is detailed in Supplemental table 3.

### Statistics

Incidence rate ratios (IRR) of all cause death were estimated using age-group stratified Poisson regression models. Estimates were adjusted for the following covariates: Sex (men/women), county of residence (categorical), calendar time (quarter and year, time varying) and medical risk group at baseline (yes/no). Each model was also stratified by sex (women/men) and calendar year (2021–2023). A separate sensitivity analysis was adjusted for age in 10-year age groups (18-29, 30-39, 40-49, 50-59, 60-69, 70-79, 80-89, ≥90) along with all other described covariates.

COVID-19 vaccination status (“unvaccinated”, “partially vaccinated” or “fully vaccinated”) was considered the exposure in all models and was handled as a time varying covariate, with each individual changing groups at the vaccination date in question.

IRRs are reported with 95% CIs, with unvaccinated person time as reference. When reporting descriptive data for time varying covariates (age group, vaccination status and calendar time) we report data from the last available point of follow-up for each individual in the given time period. Person time is reported as per 100 000 person years (py).

All analyses were conducted using Stata SE version 18.0 for Windows (StataCorp LLC, College Station, TX).

### Ethics

Beredt C19 was established under the Act on Health and Social Preparedness §2-4 to assist national and international authorities in assessing risk and implementing measures during the COVID-19 pandemic, using registry data without informed consent or need for ethical approval. National IDs were pseudonymized with an algorithm recommended by the National Security Authority, and all analyses were in compliance with the data protection impact assessment (DPIA) of Beredt C19. Only anonymous aggregated results were exported from Beredt C19. The study was approved by the Regional Committee for Medical and Health Research Ethics South-East Norway (REK South East A, ref 122745).

### Public Involvement

The study addresses a specifically stated public concern using data that was made available during the COVID-19 pandemic to evaluate such questions. However, the public at large was not directly involved in any part of the study design or conduct.

## Results

A total of 4 645 910 individuals aged ≥18 years were included during follow-up, with a mean follow-up time of 2.72 years. 3 964 864 individuals (85%) received one or more doses of COVID-19 mRNA vaccines, with 2 898 837 individuals (62%) receiving three or more doses. Vaccinated individuals were on average older than unvaccinated individuals. While 88% of individuals aged 65 years or older received three or more doses during follow-up, this was the case for only 42% among those aged 18-44 years. 20% of those aged 18-44 years did not receive any dose during follow-up, compared to only 5% among those aged 65 years or older. There was also a higher proportion of individuals with a risk condition among those vaccinated compared to those left unvaccinated through the study period (Table 1). These differences in proportion of older individuals and individuals with a risk condition between the different vaccination groups were particularly pronounced during 2021, when individuals at risk were still being prioritized for vaccination ahead of others in Norway (Supplemental table 1).

**Table 1.** Study population characteristics*.

| Sex | Age group | Variable | Period | 3+ doses | 1-2 doses | No doses |
| --- | --- | --- | --- | --- | --- | --- |
| Both | 18-44 | Total, N | 2021-2023 | 855126 | 779056 | 412538 |
|  |  | Women, N (%) | 2021-2023 | 440343 (51,5) | 355564 (45,6) | 204214 (49,5) |
|  |  | Risk group, N (%) | 2021-2023 | 101000 (11,8) | 59342 (7,6) | 20839 (5,1) |
|  | 45-64 | Total, N | 2021-2023 | 1031838 | 214211 | 206188 |
|  |  | Women, N (%) | 2021-2023 | 502309 (48,7) | 97423 (45,5) | 108443 (52,6) |
|  |  | Risk group, N (%) | 2021-2023 | 240302 (23,3) | 36508 (17,0) | 35027 (17,0) |
|  | 65+ | Total, N | 2021-2023 | 1011873 | 72760 | 62320 |
|  |  | Women, N (%) | 2021-2023 | 531013 (52,5) | 39871 (54,8) | 33540 (53,8) |
|  |  | Risk group, N (%) | 2021-2023 | 531826 (52,6) | 45717 (62,8) | 26105 (41,9) |
| Women | 18-44 | Total, N | 2021-2023 | 440343 | 355564 | 204214 |
|  |  | Risk group, N (%) | 2021-2023 | 57192 (13,0) | 30850 (8,7) | 12936 (6,3) |
|  | 45-64 | Total, N | 2021-2023 | 502309 | 97423 | 108443 |
|  |  | Risk group, N (%) | 2021-2023 | 114428 (22,8) | 17382 (17,8) | 19497 (18,0) |
|  | 65+ | Total, N | 2021-2023 | 531013 | 39871 | 33540 |
|  |  | Risk group, N (%) | 2021-2023 | 257256 (48,4) | 24456 (61,3) | 13705 (40,9) |
| Men | 18-44 | Total, N | 2021-2023 | 414783 | 423492 | 208324 |
|  |  | Risk group, N (%) | 2021-2023 | 43808 (10,6) | 28492 (6,7) | 7903 (3,8) |
|  | 45-64 | Total, N | 2021-2023 | 529529 | 116788 | 97745 |
|  |  | Risk group, N (%) | 2021-2023 | 125874 (23,8) | 19126 (16,4) | 15530 (15,9) |
|  | 65+ | Total, N | 2021-2023 | 480860 | 32889 | 28780 |
|  |  | Risk group, N (%) | 2021-2023 | 274570 (57,1) | 21261 (64,6) | 12400 (43,1) |

\*Age and vaccination status is reported at the last available point of follow-up for each individual during 2021-2023. Sex and risk group status was only registered at baseline

There was a total of 132 963 deaths during follow-up, with 116 589 (88%) occurring among those 65 years or older. In all age groups, the rate of death was lowest among those that were fully vaccinated and highest among those that were unvaccinated. The rate of death for those that were fully vaccinated compared to those that were unvaccinated was 30% lower, 27% lower and 24% lower in the age-groups 18-44 years, 45-64 years and 65 years or older (51.5 vs 73.6 per 100 000 py, 295.1 vs 405.3 per 100 000 py and 3620.2 vs 4783.8 per 100 000 py, respectively). This difference in rate of death was higher among men compared to women, particularly in the youngest age group where the difference between men and women was 19%, compared to 8% and 5% in age groups 45-64 years and 65 years or older, respectively (Table 2).

**Table 2.** Unadjusted and adjusted incidence rate ratios of death (all causes) by vaccination status.

| Sex | Age group | Period | 3+ doses |  |  | 1-2 doses |  |  | No doses |  |  |
| --- | --- | --- | --- | --- | --- | --- | --- | --- | --- | --- | --- |
|  |  |  | IRR<br>(95% CI) | aIRR<br>(95% CI)* | Deaths<br>(100 000 py<sup>§</sup>) | IRR<br>(95% CI) | aIRR<br>(95% CI)* | Deaths<br>(100 000 py<sup>§</sup>) | IRR<br>(95% CI) | aIRR<br>(95% CI)* | Deaths<br>(100 000 py<sup>§</sup>) |
| Both | 18-44 | 2021-2023 | 0,70<br>(0,64 - 0,76) | 0,42<br>(0,38 - 0,47) | 881<br>(17,10) | 0,71<br>(0,66 - 0,77) | 0,49<br>(0,45 - 0,54) | 1210<br>(23,10) | Ref | Ref | 1185<br>(16,10) |
|  | 45-64 | 2021-2023 | 0,73<br>(0,70 - 0,76) | 0,39<br>(0,37 - 0,41) | 6061<br>(20,54) | 0,80<br>(0,76 - 0,83) | 0,54<br>(0,51 - 0,57) | 3616<br>(11,22) | Ref | Ref | 3421<br>(8,44) |
|  | 65+ | 2021-2023 | 0,76<br>(0,74 - 0,77) | 0,42<br>(0,41 - 0,43) | 69545<br>(19,21) | 0,89<br>(0,87 - 0,91) | 0,77<br>(0,75 - 0,79) | 30779<br>(7,25) | Ref | Ref | 16265<br>(3,40) |
| Women | 18-44 | 2021-2023 | 0,83<br>(0,72 - 0,96) | 0,48<br>(0,40 - 0,58) | 367<br>(8,80) | 0,67<br>(0,58 - 0,77) | 0,46<br>(0,39 - 0,54) | 364<br>(10,85) | Ref | Ref | 365<br>(7,24) |
|  | 45-64 | 2021-2023 | 0,78<br>(0,73 - 0,83) | 0,41<br>(0,37 - 0,45) | 2452<br>(10,02) | 0,82<br>(0,76 - 0,88) | 0,53<br>(0,49 - 0,59) | 1372<br>(5,35) | Ref | Ref | 1196<br>(3,81) |
|  | 65+ | 2021-2023 | 0,78<br>(0,76 - 0,8) | 0,47<br>(0,45 - 0,48) | 35836<br>(10,13) | 0,93<br>(0,91 - 0,96) | 0,83<br>(0,81 - 0,86) | 16539<br>(3,91) | Ref | Ref | 8165<br>(1,80) |
| Men | 18-44 | 2021-2023 | 0,67<br>(0,60 - 0,75) | 0,40<br>(0,35 - 0,45) | 514<br>(8,30) | 0,75<br>(0,68 - 0,82) | 0,51<br>(0,46 - 0,57) | 846<br>(12,24) | Ref | Ref | 820<br>(8,86) |
|  | 45-64 | 2021-2023 | 0,71<br>(0,68 - 0,75) | 0,38<br>(0,36 - 0,41) | 3609<br>(10,52) | 0,80<br>(0,75 - 0,84) | 0,54<br>(0,50 - 0,58) | 2244<br>(5,88) | Ref | Ref | 2225<br>(4,63) |
|  | 65+ | 2021-2023 | 0,74<br>(0,72 - 0,75) | 0,37<br>(0,36 - 0,39) | 33709<br>(9,08) | 0,84<br>(0,82 - 0,87) | 0,71<br>(0,68 - 0,73) | 14240<br>(3,34) | Ref | Ref | 8100<br>(1,61) |

\*aIRR (adjusted incidence rate ratio): Adjusted for sex (man/woman), county of residence (categorical), calendar time (quarter and year) and medical risk group at baseline (yes/no)
<sup>§</sup>Person years (py)

Due to the higher proportion of individuals with a risk condition among those that received a vaccination compared to those that were unvaccinated, these differences in rates of death became more pronounced in the adjusted models. After adjusting for sex, risk condition, county of residence and calendar time the IRR of death among those that were fully vaccinated compared to those that were unvaccinated was 0.42 (95% CI 0.38-0.47) among individuals aged 18-44 years, 0.39 (95% CI 0.37-0.41) among individuals aged 45-64 years, and 0.42 (95% CI 0.41-0.43) among individuals aged 65 years or older (Figure 1). Again, this difference in rate of death between those that were fully vaccinated compared to those that were unvaccinated was higher among men than women, with a 20% lower IRR among men than women among those aged 18-44 years, compared to 8% and 27% among those aged 45-64 years and 65 years or older, respectively (Table 2).

**Figure 1.**
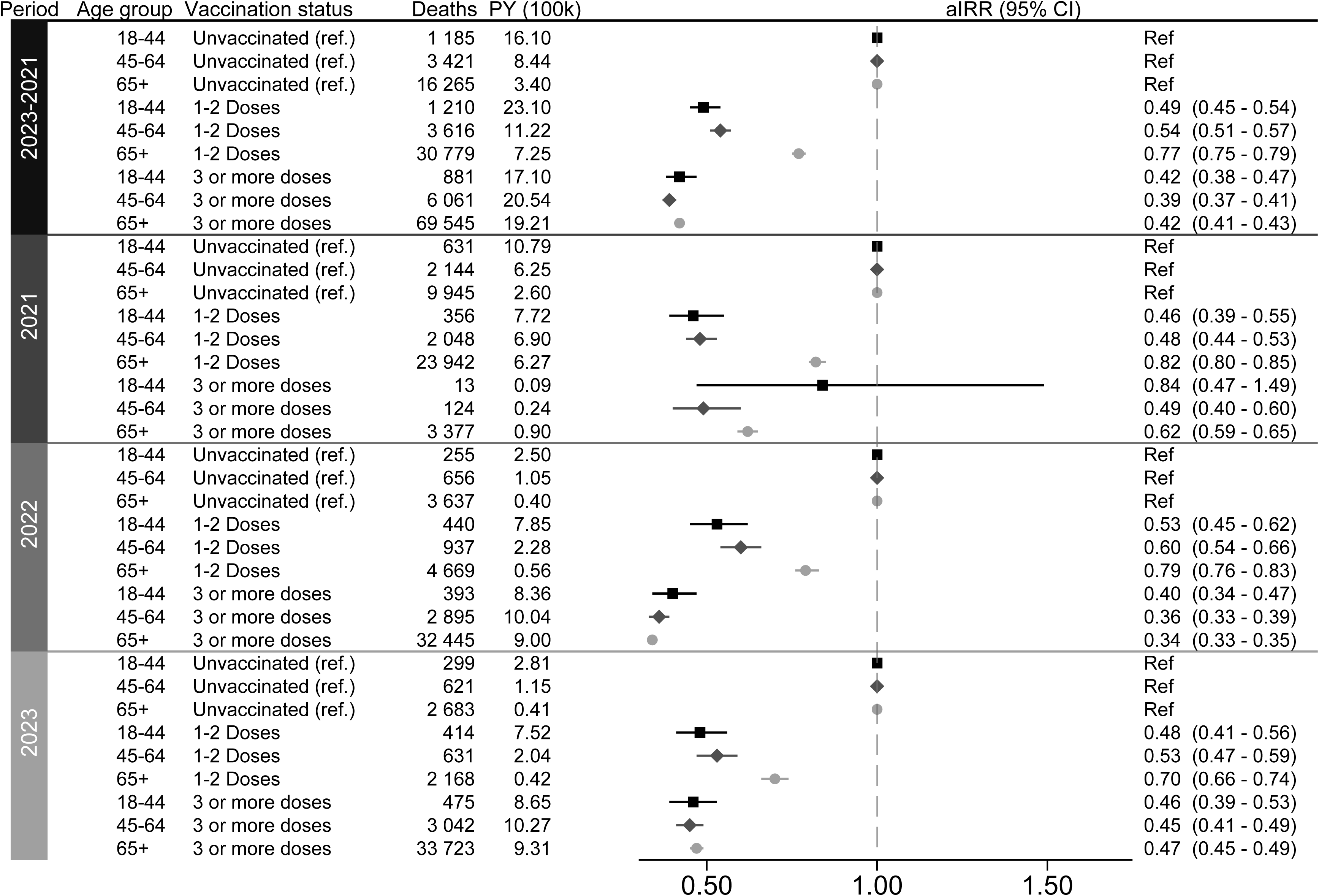
Adjusted incidence ratios (aIRR)* of death (all causes) by vaccination status *Adjusted for sex (man/woman), county of residence (categorical), calendar time (quarter and year) and medical risk group at baseline (yes/no)

The lowest IRR of death among those that were fully vaccinated compared to those that were unvaccinated occurred during 2022 for all age groups. The only subgroup of our study population that did not have a reduced risk of death among fully vaccinated compared to unvaccinated was individuals aged 18-44 years during 2021 (IRR 0.84, 95% CI 0.47-1.49). However, this estimate was based on a low number of deaths.

As a sensitivity analysis, we included the whole study population (everyone aged ≥18 years) in a separate model where we adjusted for age (categorical 10-year groups) in addition to sex, risk condition, county of residence and calendar time. In this model the fully vaccinated group had an IRR of death of 0.39 (95% CI 0.38-0.40) compared to the unvaccinated group, while those that were partially vaccinated (1-2 doses) had an IRR of 0.64 (95% CI 0.63-0.65) compared to the unvaccinated group (data not shown).

## Discussion

Our data show reduced all-cause mortality among vaccinated individuals compared to unvaccinated individuals during 2021-2023 in Norway. This difference in mortality was largest during 2022, after COVID-19 control measures were removed, and higher among men.

These results are generally in accordance with previously published data on vaccine effectiveness and safety(14). There are, however, few studies that have evaluated long-term all-cause mortality by vaccination groups on a general population level, and there have been lingering questions as to the potential impact of vaccines on the increased mortality seen during the COVID-19 pandemic(3). There will always be residual confounding in observational studies that makes it impossible to argue that the vaccine itself is the sole reason for any observed beneficial or detrimental effect, but it nonetheless seems incontrovertible that vaccinated individuals have fared better with regards to mortality risk during the covid-19 pandemic than those that remained unvaccinated.

The main strength of the current study is access to individual-level data from nationwide, high quality, mandatory health registers. Our study population comprises the whole population of Norway aged 18 years or more during the study period and includes information on covid-19 vaccinations from the mandatory national registry SYSVAK. Apart from those subject to erroneous reporting, the only vaccines not captured are vaccines received abroad and not reported to Norwegian health care personnel.

There will always be a healthy vaccinee bias present in observational studies of vaccination outcomes. We tried to address this by including information on preexisting medical conditions as well as nursing home residency to describe individuals at risk of severe outcomes. As risk group were defined at baseline, we might underestimate the total number of individuals with a risk condition. We do not expect this to have caused an overestimation of the difference in mortality rate between the vaccination groups, since we observed a higher proportion of individuals with risk conditions in the vaccinated groups. It is probably more likely that some people in the unvaccinated group are misclassified as not belonging to a risk group, due to causes for low healthcare seeking behavior such as a refugee background or low socioeconomic status(15, 16). Individuals at end-of-life receiving palliative care would also likely not receive any vaccination. However, this would mainly influence our results during the initial parts of the vaccination programme in 2021.

Vaccine effectiveness studies estimate that the protective effect of mRNA covid-19 vaccines against severe outcomes is significantly reduced six months after administration(17). In the current study we did not impose any time restriction on vaccination status, meaning that an individual remained in their designated vaccination group either until they received a vaccination that changed this, or they were censored from follow-up. This approach was chosen to address the potential existence of detrimental long-term effects of vaccination on mortality risk.

Any potential protective effect of covid-19 vaccination on all-cause mortality is likely predicated on its prevention of serious outcomes following infection. We therefore do not expect a protective effect to be present outside of time periods where there is an ongoing spread of covid-19. The number of COVID-19 cases in Norway remained low during most of 2021 until the removal of most infection control measures during the autumn, with a subsequent peak during the winter of 2021/2022 and multiple waves during 2022, which also coincided with the largest single-year drop in life expectancy in Norway since the Second World War(18, 19). There were 3 505 covid-19 associated deaths reported in Norway during 2022, compared to 965 and 1 546 in 2021 and 2023, respectively(19), and in line with this 2022 is also the part of our study period where the difference in mortality between the fully vaccinated group and the unvaccinated group is the largest. This difference in mortality risk between the vaccination groups was more pronounced among men than women, which is likely related to the increased frequency of severe outcomes following COVID-19 among men(20, 21).

We expect the differences in mortality between the vaccination groups to wane over time, as the protective effect of the vaccine itself is reduced alongside the establishment of natural immunity in the general population. The chosen study period includes the time with the highest mortality during the covid-19 pandemic in Norway but does not include a calendar year with mortality rates closer to pre-pandemic levels and an absence of widespread covid-19. This would have been interesting as a negative control with regards to differences between vaccination groups. As the population gets more booster doses, future work could also compare individuals that only received three doses to those that have regularly received additional boosters.

In conclusion, there was a reduced rate of death among individuals vaccinated with mRNA vaccines during 2021-2023 in Norway.

## Acknowledgements

The authors would like to thank Jon Michael Gran for his input regarding study design.

## Author Contributions

Dahl and Tapia had full access to all the data in the study and take responsibility for the integrity of the data and the accuracy of the data analysis.

Concept and design: Gulseth, Dahl, Tapia.

Interpretation of data: All authors.

Drafting of the manuscript: Gulseth, Dahl, Tapia.

Critical revision of the manuscript for important intellectual content: All authors.

Statistical analysis: Tapia, Dahl.

Administrative, technical, or material support: Gulseth

## Conflict of Interest

The authors report no conflict of interest.

## Funding Statement

No external funding was obtained for this study.

## Data sharing

Owing to data privacy regulations in Norway, the raw data cannot be shared. However, the data are available for research upon reasonable request to The Norwegian Institute of Public Health after approval from the Norwegian Committee for Medical and Health Research Ethics and within the framework of the Norwegian data protection legislation and any required permission from Authorities.

---

**Supplemental table 1.**
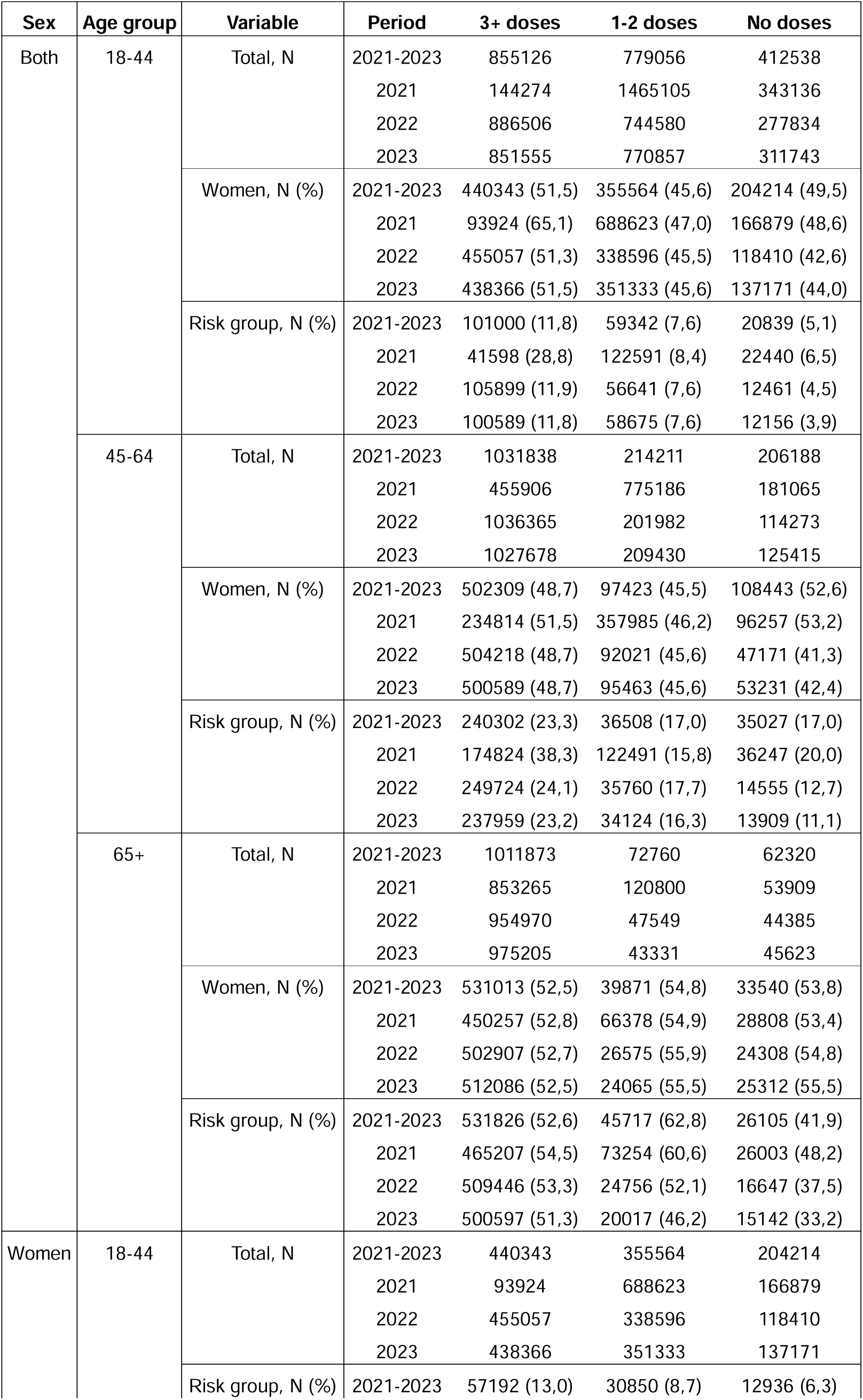
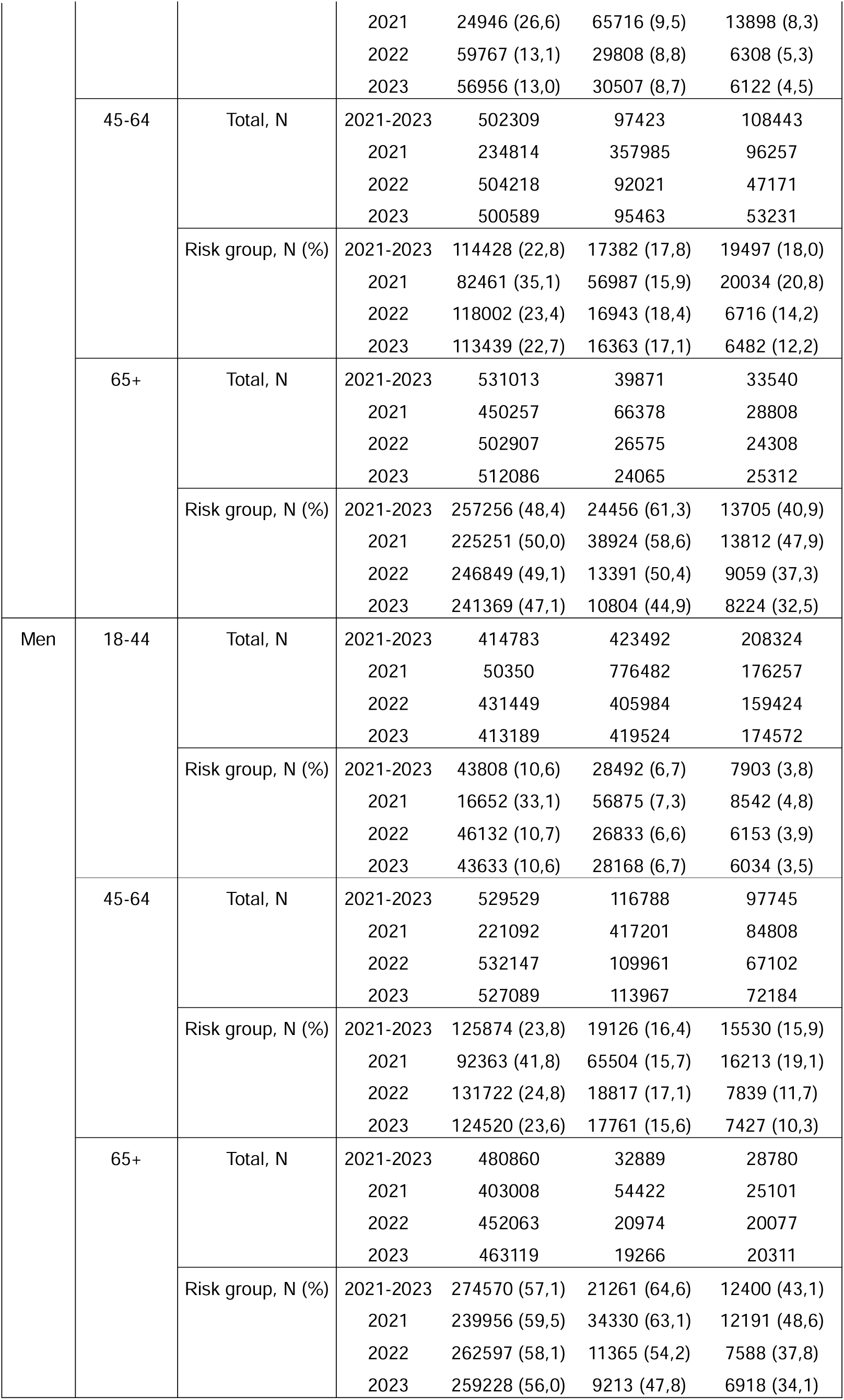
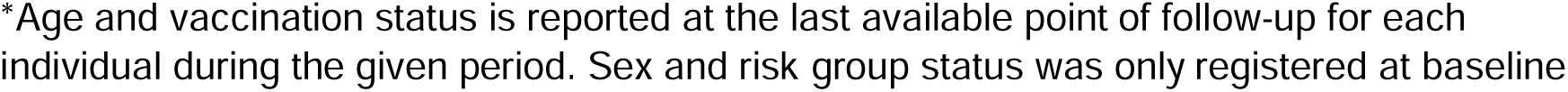
Study population characteristics by year*.

**Supplemental table 2.** Unadjusted and adjusted incidence rate ratios of death (all causes) by vaccination status and year.

| Sex | Age group | Period | 3+ doses |  |  | 1-2 doses |  |  | No doses |  |  |
| --- | --- | --- | --- | --- | --- | --- | --- | --- | --- | --- | --- |
|  |  |  | IRR<br>(95% CI) | aIRR<br>(95% CI)* | Deaths<br>(100 000 py<sup>§</sup>) | IRR<br>(95% CI) | aIRR<br>(95% CI)* | Deaths<br>(100 000 py<sup>§</sup>) | IRR<br>(95% CI) | aIRR<br>(95% CI)* | Deaths<br>(100 000 py<sup>§</sup>) |
| Both | 18-44 | 2021-2023 | 0,70<br>(0,64 - 0,76) | 0,42<br>(0,38 - 0,47) | 881<br>(17,10) | 0,71<br>(0,66 - 0,77) | 0,49<br>(0,45 - 0,54) | 1210<br>(23,10) | Ref | Ref | 1185<br>(16,10) |
|  |  | 2021 | 2,52<br>(1,45 - 4,36) | 0,84<br>(0,47 - 1,49) | 13<br>(0,09) | 0,79<br>(0,69 - 0,90) | 0,46<br>(0,39 - 0,55) | 356<br>(7,72) | Ref | Ref | 631<br>(10,79) |
|  |  | 2022 | 0,46<br>(0,39 - 0,54) | 0,40<br>(0,34 - 0,47) | 393<br>(8,36) | 0,55<br>(0,47 - 0,64) | 0,53<br>(0,45 - 0,62) | 440<br>(7,85) | Ref | Ref | 255<br>(2,50) |
|  |  | 2023 | 0,52<br>(0,45 - 0,60) | 0,46<br>(0,39 - 0,53) | 475<br>(8,65) | 0,52<br>(0,45 - 0,60) | 0,48<br>(0,41 - 0,56) | 414<br>(7,52) | Ref | Ref | 299<br>(2,81) |
|  | 45-64 | 2021-2023 | 0,73<br>(0,70 - 0,76) | 0,39<br>(0,37 - 0,41) | 6061<br>(20,54) | 0,80<br>(0,76 - 0,83) | 0,54<br>(0,51 - 0,57) | 3616<br>(11,22) | Ref | Ref | 3421<br>(8,44) |
|  |  | 2021 | 1,53<br>(1,27 - 1,83) | 0,49<br>(0,40 - 0,60) | 124<br>(0,24) | 0,86<br>(0,81 - 0,92) | 0,48<br>(0,44 - 0,53) | 2048<br>(6,90) | Ref | Ref | 2144<br>(6,25) |
|  |  | 2022 | 0,46<br>(0,42 - 0,50) | 0,36<br>(0,33 - 0,39) | 2895<br>(10,04) | 0,66<br>(0,59 - 0,73) | 0,60<br>(0,54 - 0,66) | 937<br>(2,28) | Ref | Ref | 656<br>(1,05) |
|  |  | 2023 | 0,55<br>(0,50 - 0,60) | 0,45<br>(0,41 - 0,49) | 3042<br>(10,27) | 0,57<br>(0,51 - 0,64) | 0,53<br>(0,47 - 0,59) | 631<br>(2,04) | Ref | Ref | 621<br>(1,15) |
|  | 65+ | 2021-2023 | 0,76<br>(0,74 - 0,77) | 0,42<br>(0,41 - 0,43) | 69545<br>(19,21) | 0,89<br>(0,87 - 0,91) | 0,77<br>(0,75 - 0,79) | 30779<br>(7,25) | Ref | Ref | 16265<br>(3,40) |
|  |  | 2021 | 0,98<br>(0,95 - 1,02) | 0,62<br>(0,59 - 0,65) | 3377<br>(0,90) | 1,00<br>(0,97 - 1,02) | 0,82<br>(0,80 - 0,85) | 23942<br>(6,27) | Ref | Ref | 9945<br>(2,60) |
|  |  | 2022 | 0,39<br>(0,38 - 0,41) | 0,34<br>(0,33 - 0,35) | 32445<br>(9,00) | 0,91<br>(0,87 - 0,95) | 0,79<br>(0,76 - 0,83) | 4669<br>(0,56) | Ref | Ref | 3637<br>(0,40) |
|  |  | 2023 | 0,55<br>(0,53 - 0,57) | 0,47<br>(0,45 - 0,49) | 33723<br>(9,31) | 0,78<br>(0,74 - 0,83) | 0,70<br>(0,66 - 0,74) | 2168<br>(0,42) | Ref | Ref | 2683<br>(0,41) |
| Women | 18-44 | 2021-2023 | 0,83<br>(0,72 - 0,96) | 0,48<br>(0,40 - 0,58) | 367<br>(8,80) | 0,67<br>(0,58 - 0,77) | 0,46<br>(0,39 - 0,54) | 364<br>(10,85) | Ref | Ref | 365<br>(7,24) |
|  |  | 2021 | 3,99<br>(2,11 - 7,52) | 1,20<br>(0,59 - 2,41) | 10<br>(0,06) | 0,76<br>(0,61 - 0,94) | 0,46<br>(0,34 - 0,62) | 123<br>(3,84) | Ref | Ref | 211<br>(4,98) |
|  |  | 2022 | 0,56<br>(0,43 - 0,74) | 0,45<br>(0,34 - 0,60) | 170<br>(4,30) | 0,48<br>(0,36 - 0,64) | 0,45<br>(0,33 - 0,60) | 120<br>(3,59) | Ref | Ref | 73<br>(1,04) |
|  |  | 2023 | 0,63<br>(0,49 - 0,82) | 0,50<br>(0,39 - 0,66) | 187<br>(4,44) | 0,53<br>(0,40 - 0,70) | 0,48<br>(0,36 - 0,64) | 121<br>(3,43) | Ref | Ref | 81<br>(1,22) |
|  | 45-64 | 2021-2023 | 0,78<br>(0,73 - 0,83) | 0,41<br>(0,37 - 0,45) | 2452<br>(10,02) | 0,82<br>(0,76 - 0,88) | 0,53<br>(0,49 - 0,59) | 1372<br>(5,35) | Ref | Ref | 1196<br>(3,81) |
|  |  | 2021 | 1,65<br>(1,26 - 2,17) | 0,55<br>(0,40 - 0,75) | 56<br>(0,13) | 0,94<br>(0,85 - 1,04) | 0,55<br>(0,47 - 0,63) | 826<br>(3,37) | Ref | Ref | 752<br>(2,89) |
|  |  | 2022 | 0,46<br>(0,39 - 0,53) | 0,36<br>(0,32 - 0,42) | 1141<br>(4,89) | 0,63<br>(0,53 - 0,74) | 0,58<br>(0,49 - 0,69) | 335<br>(1,04) | Ref | Ref | 220<br>(0,43) |
|  |  | 2023 | 0,54<br>(0,47 - 0,63) | 0,44<br>(0,38 - 0,51) | 1255<br>(5,00) | 0,49<br>(0,41 - 0,59) | 0,45<br>(0,37 - 0,54) | 211<br>(0,93) | Ref | Ref | 224<br>(0,49) |
|  | 65+ | 2021-2023 | 0,78<br>(0,76 - 0,8) | 0,47<br>(0,45 - 0,48) | 35836<br>(10,13) | 0,93<br>(0,91 - 0,96) | 0,83<br>(0,81 - 0,86) | 16539<br>(3,91) | Ref | Ref | 8165<br>(1,80) |
|  |  | 2021 | 1,08<br>(1,02 - 1,14) | 0,72<br>(0,67 - 0,77) | 1876<br>(0,48) | 1,05<br>(1,02 - 1,09) | 0,92<br>(0,87 - 0,96) | 12853<br>(3,36) | Ref | Ref | 4900<br>(1,35) |
|  |  | 2022 | 0,41<br>(0,40 - 0,43) | 0,38<br>(0,36 - 0,40) | 16638<br>(4,75) | 0,96<br>(0,91 - 1,02) | 0,85<br>(0,80 - 0,90) | 2531<br>(0,31) | Ref | Ref | 1844<br>(0,22) |
|  |  | 2023 | 0,57<br>(0,54 - 0,6) | 0,50<br>(0,48 - 0,53) | 17322<br>(4,90) | 0,78<br>(0,72 - 0,85) | 0,70<br>(0,65 - 0,76) | 1155<br>(0,24) | Ref | Ref | 1421<br>(0,23) |
| Men | 18-44 | 2021-2023 | 0,67<br>(0,60 - 0,75) | 0,40<br>(0,35 - 0,45) | 514<br>(8,30) | 0,75<br>(0,68 - 0,82) | 0,51<br>(0,46 - 0,57) | 846<br>(12,24) | Ref | Ref | 820<br>(8,86) |
|  |  | 2021 | 1,42<br>(0,46 - 4,43) | 0,42<br>(0,13 - 1,33) | 3<br>(0,03) | 0,83<br>(0,71 - 0,98) | 0,47<br>(0,38 - 0,58) | 233<br>(3,88) | Ref | Ref | 420<br>(5,81) |
|  |  | 2022 | 0,44<br>(0,36 - 0,53) | 0,37<br>(0,30 - 0,45) | 223<br>(4,06) | 0,60<br>(0,50 - 0,72) | 0,57<br>(0,47 - 0,68) | 320<br>(4,26) | Ref | Ref | 182<br>(1,46) |
|  |  | 2023 | 0,50<br>(0,42 - 0,60) | 0,43<br>(0,36 - 0,52) | 288<br>(4,20) | 0,52<br>(0,44 - 0,62) | 0,49<br>(0,41 - 0,58) | 293<br>(4,10) | Ref | Ref | 218<br>(1,59) |
|  | 45-64 | 2021-2023 | 0,71<br>(0,68 - 0,75) | 0,38<br>(0,36 - 0,41) | 3609<br>(10,52) | 0,80<br>(0,75 - 0,84) | 0,54<br>(0,50 - 0,58) | 2244<br>(5,88) | Ref | Ref | 2225<br>(4,63) |
|  |  | 2021 | 1,54<br>(1,21 - 1,96) | 0,46<br>(0,35 - 0,60) | 68<br>(0,11) | 0,83<br>(0,77 - 0,90) | 0,45<br>(0,40 - 0,51) | 1222<br>(3,53) | Ref | Ref | 1392<br>(3,35) |
|  |  | 2022 | 0,48<br>(0,44 - 0,54) | 0,36<br>(0,33 - 0,41) | 1754<br>(5,15) | 0,69<br>(0,61 - 0,78) | 0,61<br>(0,54 - 0,70) | 602<br>(1,24) | Ref | Ref | 436<br>(0,62) |
|  |  | 2023 | 0,57<br>(0,51 - 0,63) | 0,46<br>(0,41 - 0,51) | 1787<br>(5,27) | 0,63<br>(0,55 - 0,72) | 0,58<br>(0,51 - 0,67) | 420<br>(1,11) | Ref | Ref | 397<br>(0,66) |
|  | 65+ | 2021-2023 | 0,74<br>(0,72 - 0,75) | 0,37<br>(0,36 - 0,39) | 33709<br>(9,08) | 0,84<br>(0,82 - 0,87) | 0,71<br>(0,68 - 0,73) | 14240<br>(3,34) | Ref | Ref | 8100<br>(1,61) |
|  |  | 2021 | 0,89<br>(0,84 - 0,94) | 0,51<br>(0,47 - 0,55) | 1501<br>(0,42) | 0,94<br>(0,91 - 0,97) | 0,73<br>(0,69 - 0,76) | 11089<br>(2,91) | Ref | Ref | 5045<br>(1,24) |
|  |  | 2022 | 0,37<br>(0,35 - 0,39) | 0,31<br>(0,29 - 0,32) | 15807<br>(4,25) | 0,86<br>(0,81 - 0,92) | 0,73<br>(0,69 - 0,78) | 2138<br>(0,25) | Ref | Ref | 1793<br>(0,18) |
|  |  | 2023 | 0,54<br>(0,51 - 0,57) | 0,44<br>(0,42 - 0,47) | 16401<br>(4,41) | 0,78<br>(0,72 - 0,85) | 0,69<br>(0,63 - 0,75) | 1013<br>(0,19) | Ref | Ref | 1262<br>(0,18) |

\*aIRR (adjusted incidence rate ratio): Adjusted for sex (man/woman), county of residence (categorical), calendar time (quarter and year) and medical risk group at baseline (yes/no)
§Person years (py)

**Supplemental table 3.**
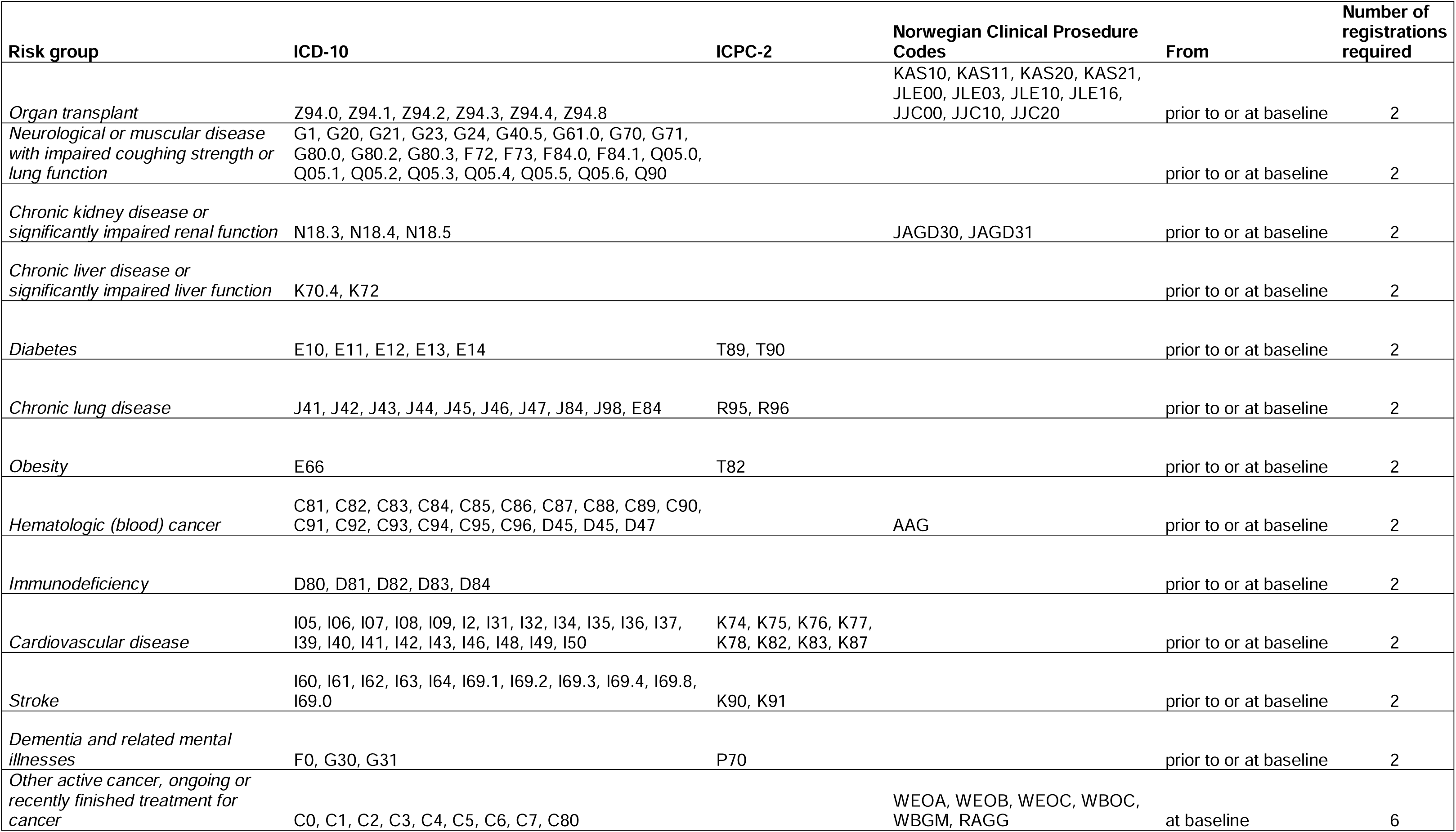
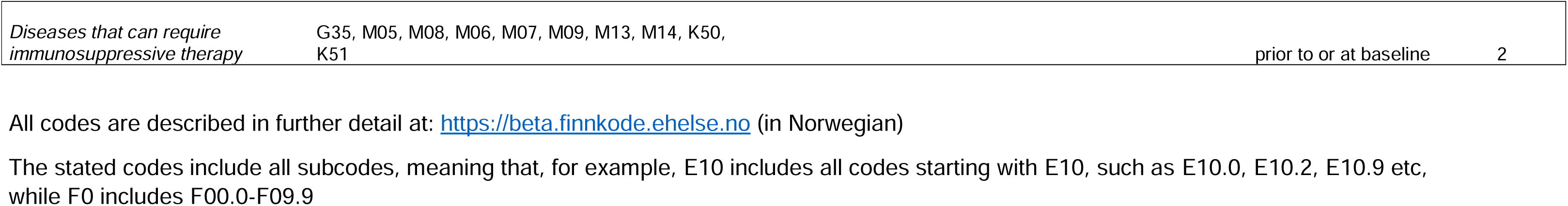
Definition of preexisting risk conditions with increased risk of severe covid-19 disease.

## Notes

### Competing Interest Statement

The authors have declared no competing interest.

### Author Declarations

The study was approved by the Regional Committee for Medical and Health Research Ethics South-East Norway (REK South East A, ref 122745)

